# Utility of Olfactory test as screening tool for COVID-19: A pilot study

**DOI:** 10.1101/2020.09.03.20187294

**Authors:** Pragyanshu Khare, Atul Munish Chander, Kanhaiya Agrawal, Satyam Singh Jayant, Soham Mukherjee, Kamalendra Yadav, Rahul Gupta, Shakun Chaudhary, Sumit Srivastava, Sanuj Muralidharan, Rijin Mohan, Shikha Chaudhary, Rimesh Pal, Sandeep Bansal, Kanthi Kiran Kondepudi, Govardhan Dutt Puri, Mahendra Bishnoi, Sanjay Kumar Bhadada

**Affiliations:** Food and Nutrition Biotechnology Division, National Agri-Food Biotechnology Institute (NABI), Knowledge city, Mohali, Punjab, India.; Department of Endocrinology, Postgraduate Institute of Medical Education and Research (PGIMER), Chandigarh, India.; Department of Rog Nidan, Shri Dhanwantry Ayurvedic College & Hospital Sector 46-B Chandigarh, India.; Department. Of Dravya Guna, Shri Dhanwantry Ayurvedic College & Hospital Sector 46-B Chandigarh, India.; Department of Kayachikitsa, Shri Dhanwantry Ayurvedic College & Hospital Sector 46-B Chandigarh, India.; Department of Anesthesia, Postgraduate Institute of Medical Education and Research (PGIMER), Chandigarh, India.

**Author notes:** Joint corresponding authors (Mahendra Bishnoi, Sanjay Kumar Bhadada). Corresponding authors: Prof. Sanjay Kumar Bhadada, Department of Endocrinology, PGIMER, Chandigarh, India., Email ID. Dr. Mahendra Bishnoi, Scientist- E, Food and Nutrition Biotechnology Division, NABI, Knowledge city, Mohali, India., Email. ID.

## Abstract

Loss of smell function (Anosmia) is reported to be associated with novel coronavirus disease 2019 (COVID-19) infection. The present study was designed to evaluate the effectiveness of an indigenously developed prototype smell test to identify/diagnose asymptomatic COVID-19 positive individuals. A panel of five different odorants belonging to Indian household with unique and mutually exclusive odor were used to develop prototype kit to test the hypothesis. The developed prototype kit was tested at 2 centers (N = 49 and 34) with slight modifications. Simultaneously, the kit was also tested on 55 (N = 35 and 20) healthy controls. Our results indicate that otherwise asymptomatic COVID-19 positive individuals were having quantifiable deficit in smell sensation. Interestingly, the variable sensitivity of different odorants was observed in different patients. None of the healthy controls reported difficulty in sensing any of the odorant, whereas, some of healthy controls did misidentify the odorants. Overall, the present study provides a preliminary data that loss in smell sensation for various odorants can be exploited as a quick and affordable screening test to identify infected cases among at risk individuals.

## 1. INTRODUCTION

Rapid spread of the Severe Acute Respiratory Syndrome Coronavirus 2 (SARS-CoV-2) and concern for viral transmission by ambulatory patients with minimal to no symptoms underline the importance of identifying early or subclinical symptoms of novel coronavirus disease 2019 (COVID-19). Asymptomatic presentation of the COVID-19 in individuals is a major challenge because they remain unidentified among the healthy population [1, 2]. Such asymptomatic carriers silently spread infections to a large proportion of normal and at risk population (with comorbidities). There are limited options to identify asymptomatic COVID-19 infected individuals [3]. Two such candidate symptoms are reported loss of smell (anosmia, 68 % of COVID-19 positive patients) and taste (ageusia, 71 % of COVID positive patients) [4].There are some studies indicating the impact of olfactory dysfunction in COVID-19 patients [5, 6], suggestive of its importance in diagnosis [7]. Available literature as well as the WHO guidelines has enumerated smell and taste loss as one of primary and initial symptoms of infection. Reduced olfactory function due to COVID-19 was also reported by some other studies [8, 9]. Till date, less is known about the olfactory function of asymptomatic COVID-19 individuals. The present study was designed to evaluate the effectiveness of an indigenously developed prototype smell test to identify/diagnose asymptomatic COVID-19 positive individuals. For this purpose, the characteristic smell test derived from the herbs/spices commonly present in Indian kitchens were deployed to develop a five-minute odor test. Given the non-availability/expensive nature of testing kits, this test may enable us to perform rapid and wider testing along with its use as safety measures to identify COVID-19 patients. In addition to this, the test has a potential to be one of the preliminary scanning method along with infrared thermometry at the entry points of hospitals, government and private offices, shops and other places of public dealing in order to have a safe cordon.

## 2. METHODOLOGY

The study was approved by Institute Ethics committee, Post Graduate Institute of Medical Education and Research (PGIMER), Chandigarh. Trial was registered in Clinical Trial Registry of India with CTRI Reg. No. CTRI/2020/05/025487 (http://ctri.nic.in).

The confirmed adult patients diagnosed (as family members, primary and secondary contacts of symptomatic COVID-19 infected individuals) positive by RT-PCR test for COVID-19 infection were recruited from the COVID19 ward of PGIMER, Chandigarh (N = 49) and associated Hospital (COVID-19 ward at Shri Dhanwantary Ayurvedic College, Sector 46, Chandigarh (N = 35). COVID-19 positive asymptomatic patients of any age/ sex were included in this study. Moreover, apparently healthy subjects were recruited although were not tested for the COVID-19 confirmatory test. Patients on a regular medication with probability of loss of smell and taste sensation was not included in the study. Smokers and those addicted to chewing paan and similar materials were also excluded because their smell sensation is already suspected to be affected.

In total, 25 odorants were screened for their potential to be a candidate and its effect on desensitization of smell sensation (Fig. 1A). We graded these odorants in their potency and their familiarity to the population. Based on our online survey (100 participants), we have selected 5 odorants which fits into our selection criterion and represents different smell categories with varied strength. Those 5 odorants with maximum familiarity among these 100 participants were selected to test in the trial. These 5 odorants were, coconut oil (O1), cardamom (O2), fennel (O3), peppermint (O4), and garlic (O5). Based on these we developed a prototype test kit. These odorants were filled in sterile 2 ml tubes in the crushed form (cardamom and fennel) or directly oil/chemical form (oil in case of coconut oil, menthol for peppermint and Di-allyl sulphide for garlic). The tubes were packed in one bag along with the response sheet carrying a questionnaire about whether an individual can smell the odorants in tube followed by identification of the smell if answered yes. Multiple options were given to make it easier for the participants. The first study was performed in 49 asymptomatic (no comorbidities) COVID19 positive individuals admitted at PGIMER, Chandigarh. Simultaneously, 35 control individuals also undertook the test. Following first study, we modified the test to include (a) blank (water) smell and (b) change the order of each odorant in each kit and (c) test was performed in double blind form. All these changes were made to confirm the findings of first study. Second study was performed at The Dhanwantry Ayurveda Hospital, Sector 46, Chandigarh. A study was performed on 35 asymptomatic or mild symptoms (no comorbidities) COVID 19 positive individuals admitted at COVID facility. Simultaneously, 25 control individuals also undertook the test.

**Figure 1.**
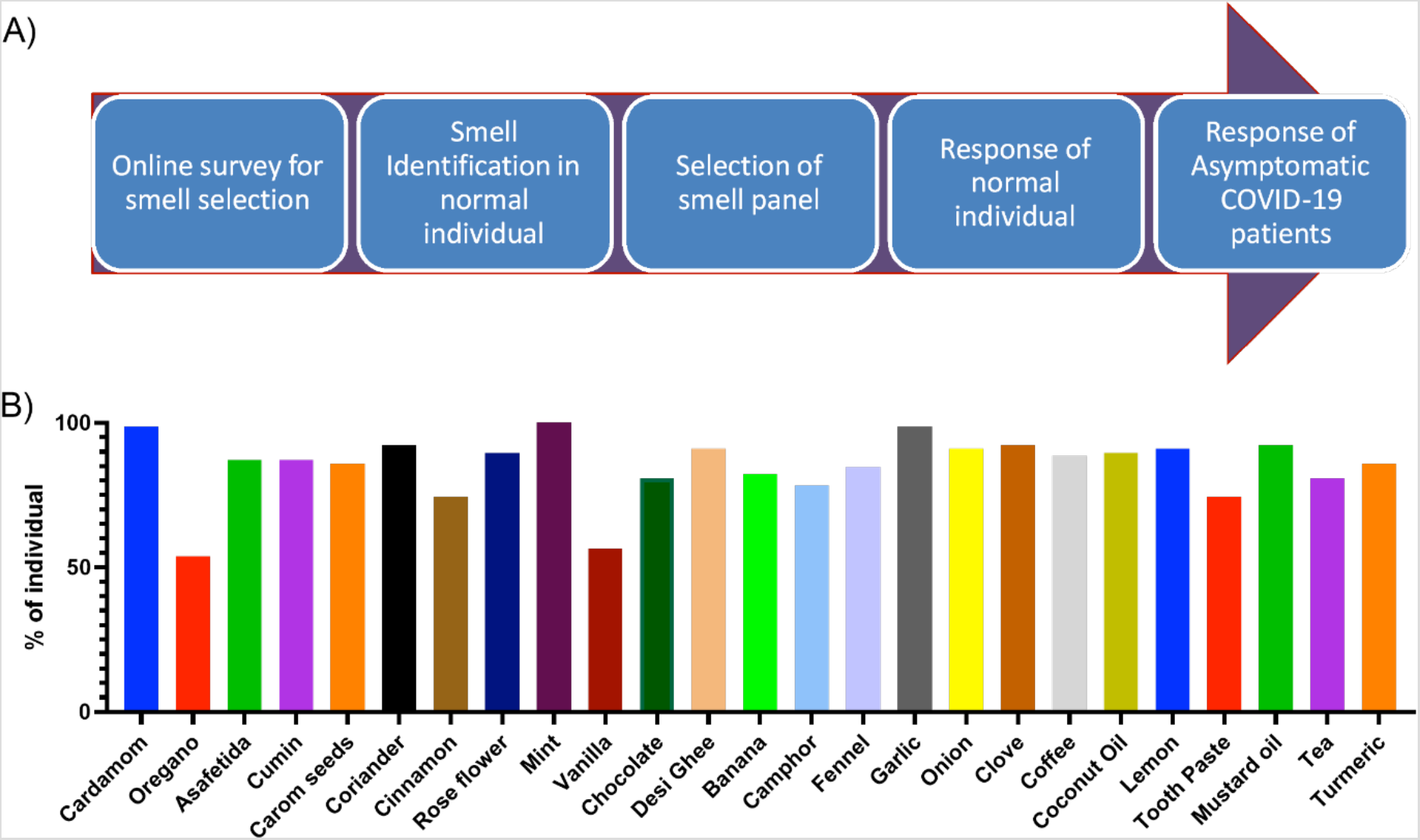
A) Schematic diagram showing experiment design and B) Preference and familiarity of individuals to the common household odorants on the basis of online survey.

## 3. RESULTS AND DISCUSSION

Fig1A shows the overall plan of experiment in stepwise manner. Fig1B shows the preference and familiarity of individuals to the common household odorants. We selected 5 of these based on maximum familiarity, different types and strength of smells. As described in Fig2A and B, in study 1 (at COVID-19 facility PGIMER) COVID-19 asymptomatic positive individuals has significant loss of smell sensation. The two types of outcomes were received while odor test. In both the groups (COVID-19 patients and healthy controls), there were some respondents who could sense smell but were unable to recognize it. The second type of respondents were completely unable to sense a smell, were only noticed in COVID-19 group. Similar to earlier reports we also observed smell deficits in nearly 50% of COVID-19 patients [10]. Fig2A is a cumulative data of % COVID-19 positive individuals who were unable to either smell or identify the odorants correctly, or both, which goes as high as 48.9 % and 22.5% for single and two odorants respectively (Fig2A). This data is clearly showing that smell deficit is not a global smell perception loss but depends on the type of odorant, mechanism of which is unknown yet. In the same line, we observed that, only 6.1 % COVID-19 positive individuals were not able to smell/identify all 5 odorants (Fig2A). Furthermore, the difference in identification of smell and complete inability to sense the smell was different among COVID-19 positive individuals as 38.8% and 16% individuals were completely unable to smell at least single or two odorants respectively. In the present study we observed that 4.1 % individuals were not able to smell any of the 5 odorants (Fig2B) used in the study. Interestingly, no healthy individuals were completely unable to sense any of the odorant however, 14 % healthy individual misidentified at least one odorant (Fig2A, B). Further, we tend to identify whether there is any specific smell (out of these odorants) which these individuals are not able to smell. Highest misidentification was observed in peppermint (O4, 36.7%) and coconut (O1, 22.4%) smells respectively. Similarly, highest non response (unable to smell) was also observed in peppermint (O4, 24.5%) and coconut (O1, 20.4%) smells in COVID-19 positive individuals suggesting that these are specific smells more compromised during COVID-19 infection (Fig2C). During our first study we noticed that being asymptomatic and known to each other (as asymptomatic individuals were either primary or secondary contacts of symptomatic individuals), there is a chance of interaction and discussions among individuals which can affect the study data. Hence, we made few changes in our next study protocol and modified the test to include (a) blank (water) and (b) change the order of each odorant in each kit and (c) test was performed in double blind (hospital caretaker and patient were unaware) form. Following study 2, Fig2D is showing the cumulative data of % COVID-19 positive individuals who were unable to either smell or identify the odorants correctly, or both which goes as high as 85.3 % and 61.8% individuals for single and two odorants respectively and 20.6% individuals were not able to smell/identify all 5 odorants. Similar to the first study, 29.4 % and 8.9% COVID-19 positive individuals were completely unable to sense at least one and two odorants respectively (Fig2E). In this study 20% and 5% healthy individual misidentified one and two odorants respectively however, similar to our previous study no healthy individual was completely unable to sense any of the odorants. From both our studies it was clear that there is some degree of smell loss in these individuals. This study has limitation to not able in acquisition of follow-up data of these anosmic/ hyposmic patients at recovery from disease, because the study was planned with a motive of commercialization rather than to study the pathophysiology of anosmia in infection.

**Figure 2.**
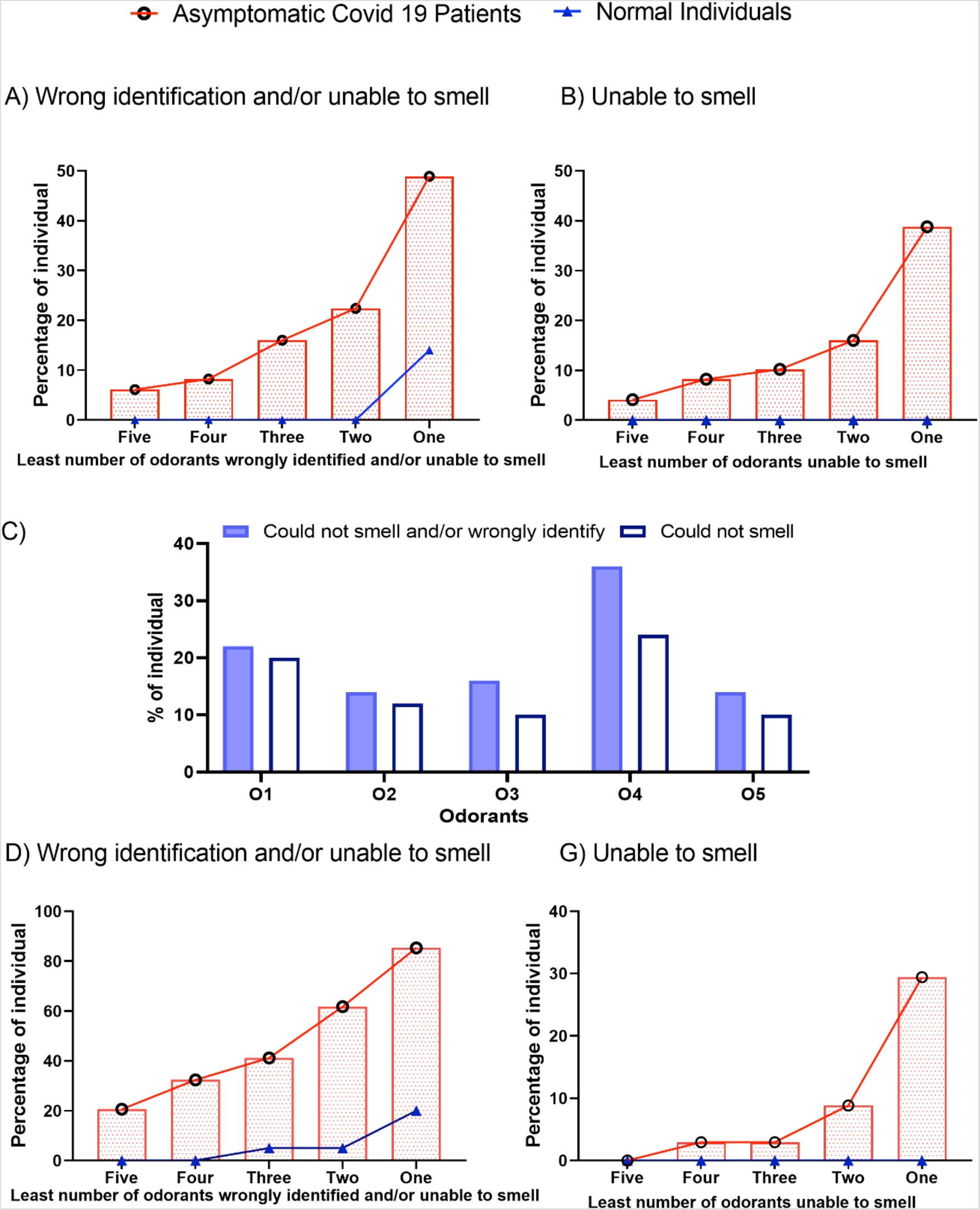
A) Percentage of individual misidentified or unable to smell (responded no smell) least number of smells in trail 1, B)- Percentage of individual completely unable to smell (responded no smell) least number of smells in trail 1; C)- Odorant based misidentification and complete inability to specific smells in trial1; D)- Percentage of individual misidentified or unable to smell (responded no smell) least number of smells in trail 2, D)- Percentage of individual completely unable to smell (responded no smell) least number of smells in trail 2.

In our study, although, the COVID-19 asymptomatic patient group was not presenting symptoms associated with congestion or nasal blockage but still the traits of loss of smell were noticed in the respondents indicating the importance of mechanisms other than congestion in pathoetiology of COVID-19 associated anosmia [6, 11]. Moreover, the variable response of asymptomatic patient to the different types of odors can be attributed to the level of virus induced dysfunctions leading to neuronal damage due to COVID-19 infection which is not completely known yet but a recent study has pointed towards this [12]

This type of tests if developed and validated in larger set of population will have many advantages. It will enable us do rapid and wider testing to identify at risk individuals as well localities. Also, it allows independent non-invasive self-testing facility. This test has a potential to be one of the scanning/screening tests along with temperature sensation (thermometer) at the entry points of hospitals, government and private offices, shops and other places of public dealing in order to have a safe cordon.

In summary, the present study is a step forward in exploiting hyposmia and anosmia in asymptomatic COVID-19 patients by using indigenous odorants. To the best of our knowledge, this is the first prominent study in India, which is not based on solely on online questionnaires or telephonic interviews, where a dedicated panel of 5 odorants to which the patients have responded and filled in their responses have been used. Responsiveness of each COVID-19 patient to different odorant was different as they were not sensitive to all the smells or viceversa. Therefore, a large and diverse panel of multiple odorants can be utilized for the screening purpose with better sensitivity. Given that testing is the key, this cost effective test gives us possibility to screen each individual in India, which is unless not possible through any other form of testing. This smell test can be used as a swift-tool for community survey to detect high risk pockets in a large population. The odor test will enable self-reporting, early identification and isolation of asymptomatic/ pre-symptomatic cases or patients with mild symptoms. Thus, the odor test will facilitate targeted mobilization of resources for tracing, testing, and isolation of cases in these areas.

## Data Availability

Relevant data is included in the article. There is no such data from this work that need submission in repositories.

